# Monitoring of COVID-19 Pandemic-related Psychopathology using Machine Learning

**DOI:** 10.1101/2021.07.13.21259962

**Authors:** Kenneth C. Enevoldsen, Andreas A. Danielsen, Christopher Rohde, Oskar H. Jefsen, Kristoffer L. Nielbo, Søren D. Østergaard

**Affiliations:** Center for Humanities Computing Aarhus, Aarhus University, Aarhus, Denmark; Department of Affective Disorders, Aarhus University Hospital - Psychiatry, Aarhus, Denmark; Department of Clinical Medicine, Aarhus University, Aarhus, Denmark

**Author notes:** Corresponding author: Kenneth C. Enevoldsen, Center for Humanities Computing Aarhus, Aarhus University, Jens Chr. Skous Vej 4, 8200 Aarhus, Denmark.

**Keywords:** Coronavirus, COVID-19, Pandemic, Mental Disorders, Machine Learning, Natural Language Processing

## Abstract

The COVID-19 pandemic has been shown to have major impact on global mental health. We show that the developments in COVID-19 pandemic-related psychopathology can be meaningfully monitored using machine learning methods providing invaluable insights to psychiatric services and of the management of the psychological consequences of the COVID-19 pandemic. The COVID-19 pandemic-related psychopathology among patients with mental illness was found to covary with the pandemic pressure, however, was less pronounced during the second wave compared to the first wave of the pandemic - possibly due to habituation.

---

The COVID-19 pandemic is believed to have a major negative impact on global mental health due to the viral disorder itself as well as the lockdowns, social distancing, isolation, fear, and increased uncertainty associated with the pandemic (Brooks et al., 2020; Sønderskov et al., 2021; Szcześniak et al., 2021). Individuals with preexisting mental illness are likely to be particularly vulnerable to the psychological stress associated with the COVID-19 pandemic (Jefsen et al., 2020; Rohde et al., 2020). Indeed, based on screening of clinical notes from electronic health records, we recently showed that many patients with mental illness appeared to develop COVID-19 pandemic-related psychopathology – i.e., symptoms of mental illness that seemed to be either directly or indirectly caused by the pandemic – during the first wave of the pandemic in the spring of 2020 (Rohde et al., 2020). Here, we continue this effort in an attempt to monitor the development of COVID-19 pandemic-related psychopathology over the extended course of the pandemic.

Based on the clinical notes assessed for COVID-19 pandemic-related psychopathology by Rohde et al., (2020), two types of supervised machine learning models were trained to classify notes as “pandemic-related psychopathology” and “not pandemic-related psychopathology”. Rohde et al. (2020) manually screened a total of 11 072 clinical notes from the electronic health record system of the Psychiatric Services of the Central Denmark Region from the period from February 1^st^ to March 23^rd^, 2020 (the first confirmed case of COVID-19 in Denmark was on February 27^th^ 2020). The 11 072 clinical notes were extracted from all 412 804 clinical notes from the period in question using COVID-19 related search queries: “corona” (corona), “covid” (covid), “virus” (virus), “epidemi” (epidemic), “pandemi” (pandemic), “smitte” (contaminate/contamination), including compound words. The 11 072 notes were labeled manually with regard to COVID-19 pandemic-related psychopathology by CR and OHJ with discrepancies resolved through discussion and consultation with SDØ. 1 357 of the 11 072 clinical notes described COVID-19 pandemic-related psychopathology. For more details, please see Rohde et al. (2020).

Using the 11 072 labeled clinical notes (1357 describing COVID-19 pandemic-related psychopathology and 9715 without COVID-19 pandemic-related psychopathology), two types of machine learning models (XGBoost and Naïve Bayes) were trained using a 5-fold cross-validated grid search on the training set. The training set consisted of 70% of the 11 072 clinical notes (randomly drawn). The models were trained on the natural text in the clinical notes, the text field indicator (e.g. ‘observations of the patient’, ‘current mental status’, ‘plan’, ‘conclusion’) patient’s sex and diagnosis, and inpatient or outpatient status. Subsequently, the trained models were validated on a test set consisting of the remaining 30% of the clinical notes. The best performing model was then applied to the clinical notes (selected using the same COVID-19 related search queries as in Rohde et al. (2020)) from the period from April 1^st^, 2020 to March 23^rd^, 2021. These notes were labeled dichotomously (pandemic-related psychopathology; yes/no) based on a threshold calculated from the test set to result in 95% specificity. Finally, to validate model performance in this later period, 500 clinical notes from this period were manually labeled by CR and OHJ (similar approach as that used by Rohde et. al. (2020)), and compared to the labeling performed by the model. For more information on model selection, preprocessing, and validation, see the Supplementary Material. This project was approved by the Chief Medical Officer of Psychiatry in the Central Danish Region as part of quality development aimed at monitoring the level of pandemic-related psychopathology during the COVID-19 pandemic.

The best performing model was XGBoost (see the Supplementary Material), which achieved an area under the receiver operating characteristic curve (AUC) of 0.88 on the test set. The results obtained when applying the XGBoost model to the clinical notes from the period from April 1^st^, 2020 to March 23^rd^, 2021 are available in Figure 1, which shows the development in pandemic-related psychopathology alongside the number of deaths due to COVID-19 in Denmark. There was a positive correlation between the level of pandemic-related psychopathology and the number of deaths due to COVID-19 in Denmark (*R*^2^ 0.13, 95% *CI* [0.06, 0.20], p < 0.0001, smoothed *R*^2^ 0.20, 95% *CI* [0.12, 0.29], *p* < 0.0001). A total of 78 540 clinical notes were labeled using the model of which 7125 was found to describe pandemic-related psychopathology. The validation of the model revealed a slight performance decrease to AUC=0.83, but no systematic decline in performance over time (see Figure S1 in the Supplementary Material) as might have been expected if there was a distribution shift, i.e. a substantial change in the content of the clinical notes over time.

**Figure 1:**
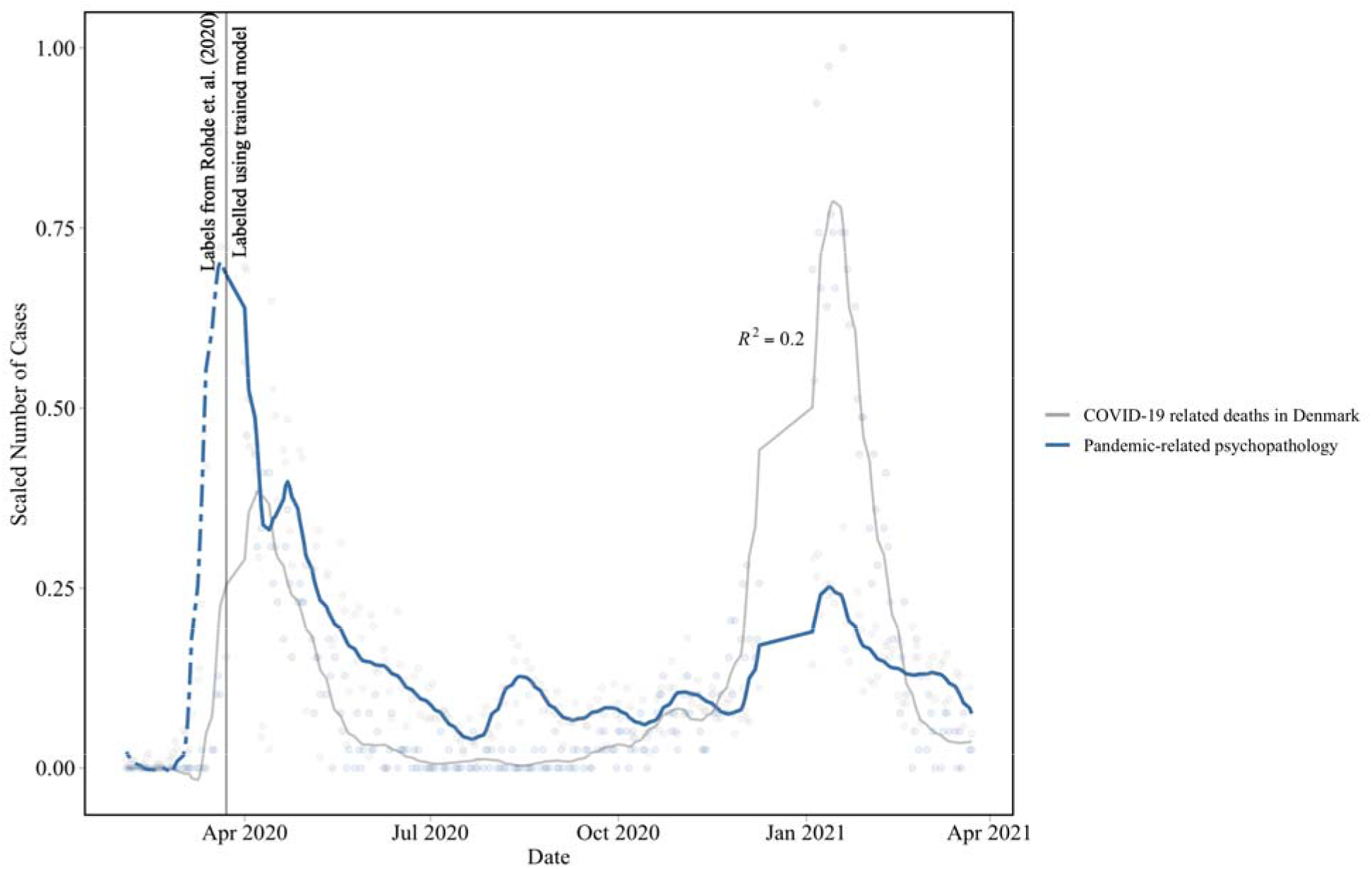
COVID-19 pandemic-related psychopathology and deaths due to COVID-19 over time. Both signals are smoothed using a local polynomial regression with an and holidays and weekends are excluded. The scale is normalized between 0 and 1 to allow for comparison. The dotted line indicates the transition from the clinical notes labeled by Rohde et al. (2020) to the trained model. Data points representing less than 5 cases of pandemic-related psychopathologies are not visualized.

The correlation between the number of clinical notes describing pandemic-related psychopathology and the number of deaths due to COVID-19 in Denmark indicates that the mental health impact of the COVID-19 pandemic among patients with mental illness covaries with the pandemic pressure. This finding is clinically meaningful and compatible with the covariation between the pandemic pressure and psychological well-being observed at the general population level in Denmark (Sønderskov et al., 2021). The correlation, however, seems to be less pronounced for the second wave compared to the first wave of the pandemic. As there was no systematic decline in model performance over the observation period, the weakened correlation could likely be due to habituation – i.e. that patients are less sensitive to the development in the pandemic as time passes, possibly because the situation appears less insecure compared to the initial phase of the pandemic. Both the correlation in itself and its weakening over time are important observations that may aid psychiatric services in the planning of the management of the psychological consequences of the COVID-19 pandemic. Furthermore, the results are testament to the potential of applying machine learning on structured and natural text data from electronic health records in clinical psychiatry.

## Data Availability

Due to the sensitive nature of the data, they are only available for quality development projects to employees in the Central Denmark Region upon application to, and approval by, the Chief Medical Officer of Psychiatry.

## Acknowledgements

The authors thank Bettina Nørremark from the Business Intelligence Office in the Central Denmark Region for her assistance with extraction of data.

