## Supplementary material for "Monitoring of COVID-19 Pandemic-related Psychopathology using Machine Learning"

**Model and parameter search**

Table S1 lists the included models and their respective hyperparameters. Table S2 lists the preprocessing hyperparameters. All hyperparameters were searched using a grid. Hyperparameter values in bold were used for fitting the final model. Note that Naïve Bayes provides a strong baseline model and that optimization over only a few key learning algorithms is often preferable as it limits the search space and allows for easier validation, maintenance, and explainability.

Table S1: Model and their hyperparameters

| **Model** | **Hyperparameters** | **Potential Values** | **Highest obtained test AUC** |
| --- | --- | --- | --- |
| XGBoost | Learning rate | 0.1, 0.2, **0.3** | 0.8875 |
|  | Number of estimators | 50, 100, **200** |  |
|  | Macimum depth | **3**, 6, 12, 18 |  |
|  | Minimum child weight | 1, 2, **3** |  |
|  | Gamma | 0.0, **0.1**, 0.2 |  |
| Naïve Bayes | N/A | N/A | 0.8319 |

*Values in bold mark those achieving the highest AUC.* N/A: not applicable.

Table S2: Preprocessing hyperparameters

| **Hyperparameters** | **Potential Values** |
| --- | --- |
| Transformation | **TF-IDF** |
| N-gram range | (1, 2) |
|  | (1, 3) |
|  | **(1, 4)** |
| Minimum number of words | 2 |
|  | **5** |

*Values in bold mark those achieving the highest AUC.*

N-grams which appeared in more than 50% of the documents were removed as these are unlikely to be discriminative. Similarly, hapaxes (words which only appear once), were removed as the estimated effect of these variables is very uncertain. Before validating the model in the test set we also experimented with over and under sampling using e.g. Synthetic Minority Oversampling Technique (SMOTE), but found no effect of this for the XGBoost model using cross-validation, However the Naive Bayes model obtained a slightly higher performance (AUC=0.85) using SMOTE.

Model training and selection was performed in Python (version 3.7.9) using scikit-learn (Buitinck et al., 2013, version 0.23.2), and imblearn (Lemaître et al., 2017, version 0.7.0). For XGBoost the python package xgboost was used (Chen & Guestrin, 2016, version 1.0.2).

**Model application and validation**

Besides cross-validation during training and a validation during testing, the trained XGBoost model was also validated on a random sample of 500 clinical notes over the period in which the model was applied to (April 1st 2020 to March 23rd 2021). This was done to investigate if the performance of the model changed over time. These clinical notes were extracted and labeled manually by CR and OHJ using an approach similar to that reported by Rohde et al. (2020) – i.e. CR and OHJ both labeled all 500 notes and consensus was reached after discussion of discrepant labellings. Based on the test set, a threshold (0.33) was set such that the model obtained a 95% specificity and consequently a 36% sensitivity. This threshold was used to label notes from April 1st 2020 to March 23rd 2021. When comparing the model labelling of the 500 additional notes with the labelling performed by CR and OHJ (consensus between the two), we found a minor performance decrease (92% specificity and 33% sensitivity), but no systematic decline in performance over time (see Figure S1).

Figure S1. Histogram showing the total number of random samples (grey) and the total number of misclassifications (red).

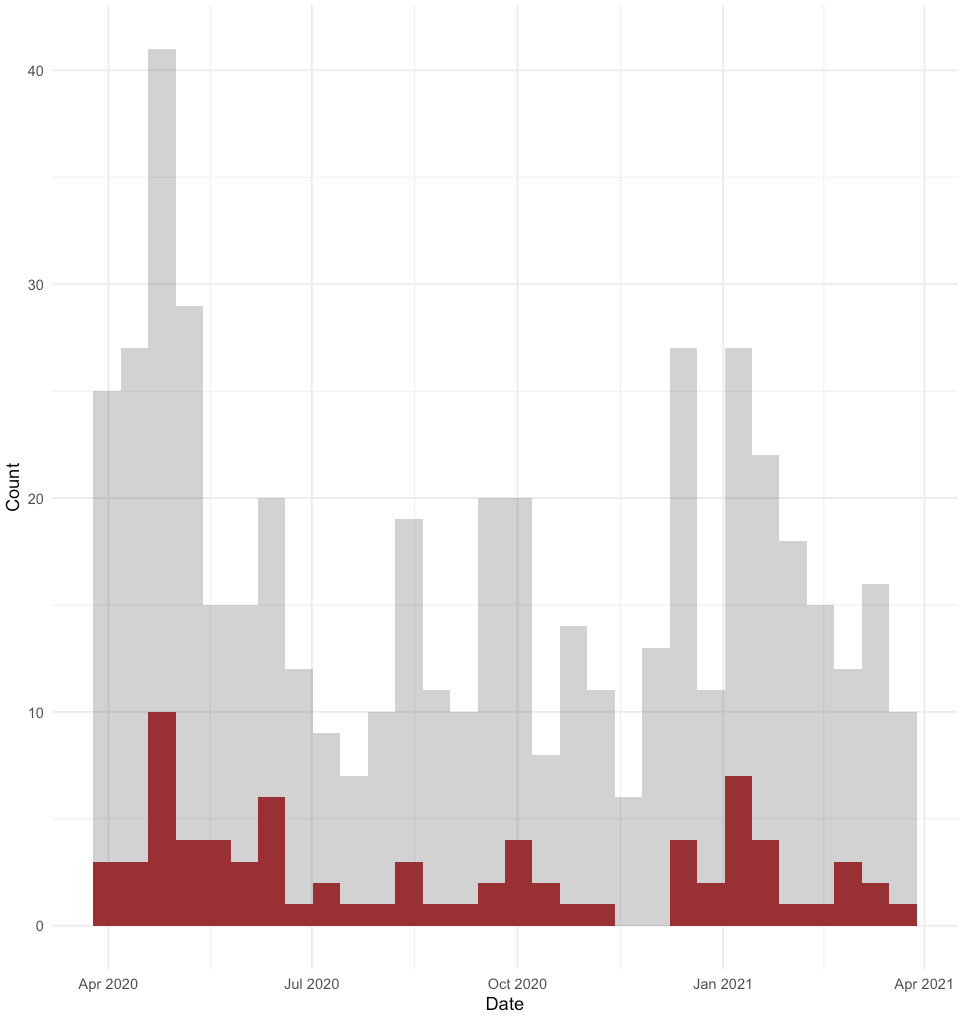
